# Mendelian randomisation with proxy exposures: challenges and opportunities

**DOI:** 10.1101/2024.10.21.24315891

**Authors:** Ida Rahu, Ralf Tambets, Eric B. Fauman, Kaur Alasoo

## Abstract

A key challenge in human genetics is the discovery of modifiable causal risk factors for complex traits and diseases. Mendelian randomisation (MR) using molecular traits as exposures is a particularly promising approach for identifying such risk factors. Despite early successes with the application of MR to biomarkers such as low-density lipoprotein cholesterol and C-reactive protein, recent studies have revealed a more nuanced picture, with widespread horizontal pleiotropy. Using data from the UK Biobank, we illustrate the issue of horizontal pleiotropy with two case studies, one involving glycolysis and the other involving vitamin D synthesis. We demonstrate that, although the measured metabolites (pyruvate or histidine, respectively) do not have a direct causal effect on the outcomes of interest (red blood cell count or vitamin D level), we can still use variant effects on these downstream metabolites to infer how they perturb protein function in different gene regions. This allows us to use variant effects on metabolite levels as proxy exposures in a *cis*-MR framework, thus rediscovering the causal roles of histidine ammonia lyase (*HAL*) in vitamin D synthesis and glycolysis pathway in red blood cell survival. We also highlight the assumptions that need to be satisfied for *cis*-MR with proxy exposures to yield valid inferences and discuss the practical challenges of meeting these assumptions.

## Introduction

A key challenge in human genetics is identifying modifiable causal risk factors for complex traits and distinguishing those from other biomarkers with no causal effect. For example, many cardiovascular disease loci are also associated with low-density lipoprotein (LDL) cholesterol level, a known causal risk factor for cardiovascular disease (Ference et al., 2012; Richardson et al., 2022). Furthermore, Mendelian randomisation (MR) studies have demonstrated that individuals with a genetic predisposition to lower LDL cholesterol level also have a reduced risk of cardiovascular disease (Voight et al., 2012). This link has been confirmed by clinical trials demonstrating the success of lipid-lowering therapies in reducing cardiovascular disease risk (Mihaylova et al., 2024). In contrast, results from MR studies are not consistent with a causal link between C-reactive protein (CRP) and cardiovascular disease, despite a strong observational correlation (C Reactive Protein Coronary Heart Disease Genetics Collaboration (CCGC) et al., 2011).

The hope of replicating the success in identifying modifiable biomarkers like LDL cholesterol for other traits has prompted the high-throughput measurements of thousands of accessible molecular traits from tens to hundreds of thousands of individuals in existing large biobanks. These molecular traits include plasma metabolites (Karjalainen et al., 2024; Richardson et al., 2022; Smith et al., 2022), plasma proteomics (Sun et al., 2018, 2023), and transcriptomic data from whole blood (Võsa et al., 2021). Relying on easily accessible whole blood and plasma samples has enabled these studies to attain sufficient sample sizes to capture associations with low-frequency variants, as well as genetic associations with small effects. As a result, these studies now routinely identify thousands of associations. Furthermore, while early proteomic and transcriptomic studies focused on genetic variants located near the protein-coding genes to map *cis* quantitative trait loci (*cis*-QTLs, Figure 1A), increased sample sizes mean that most detected associations are now located in *trans* and affect the target gene or protein levels via the activity of *trans*-acting factors (typically other proteins, Figure 1C) (Sun et al., 2023). These genetic resources provide a large number of genetic instruments for MR studies, contributing to the rapid increase in MR studies in the literature (Richmond & Davey Smith, 2022; Sanderson et al., 2022; Stender et al., 2024).

**Figure 1.**
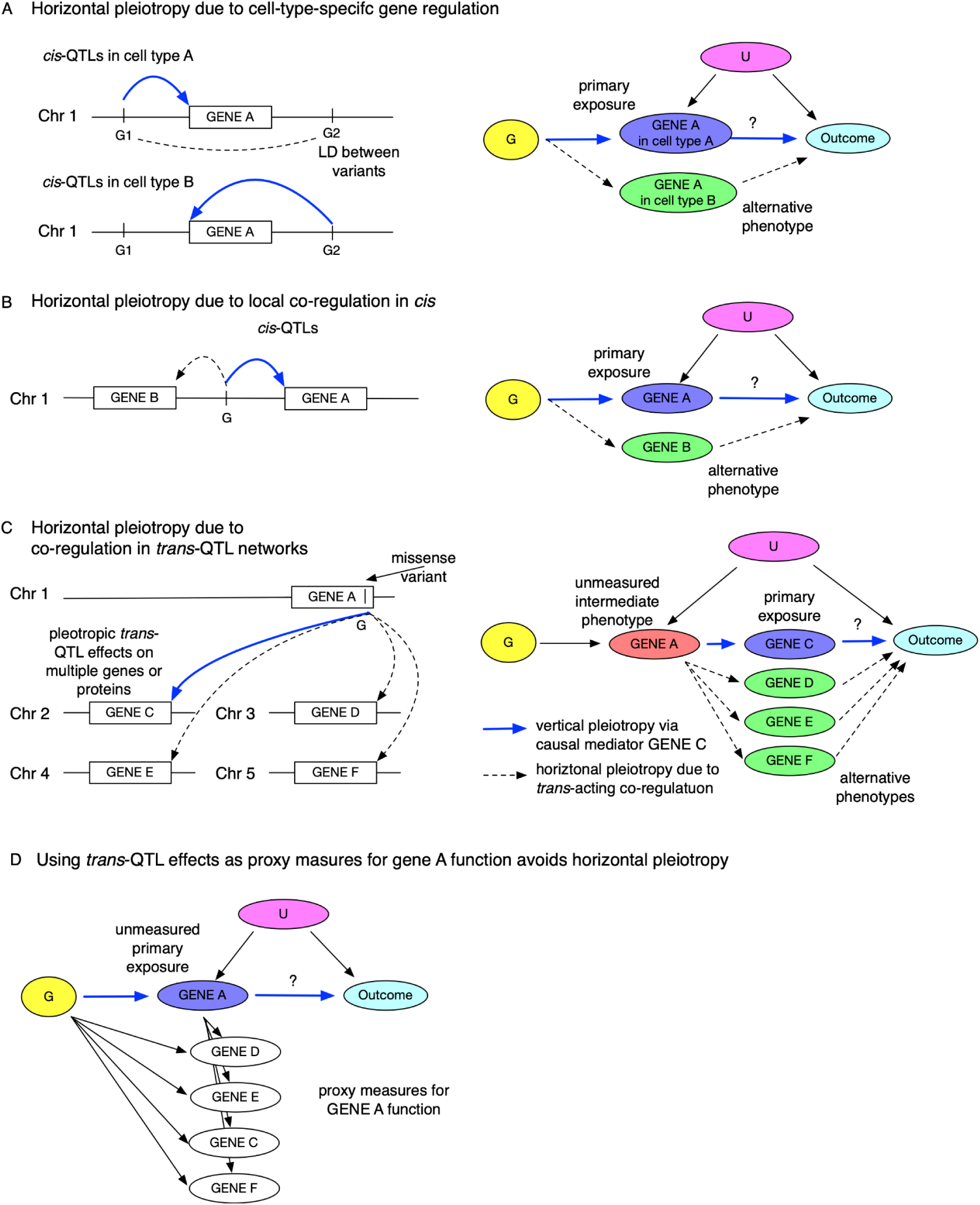
Some molecular mechanisms of horizontal pleiotropy. (**A**) In the presence of cell-type-specific QTLs, the same gene profiled in a different cell type can be a source of horizontal pleiotropy. Note that cell-type-specific QTLs G1 and G2 could be in linkage disequilibrium (LD) with each other (dashed line), further complicating the inference. (**B**) Horizontal pleiotropy due to local co-regulation of gene expression. Example of a *cis*-QTL variant G that is associated with the expression of gene A (primary exposure) and also with a neighbouring gene B (alternative phenotype). In the causal diagram, potential horizontal pleiotropy is limited to a small number of locally co-regulated genes. (**C**) Horizontal pleiotropy due to co-regulation in *trans*-QTL networks. Example of a *trans*-QTL variant G that is associated with the expression of genes C−F on different chromosomes. Note that the *trans*-QTL effect of variant G on genes C−F is typically mediated by at least one gene in the *cis* region (e.g., gene A). High degree of horizontal pleiotropy can make it challenging to identify the true causal mediators. The variant effect on *cis* gene A function is treated as an unmeasured intermediate phenotype (vertical pleiotropy) (**C**) The same scenario as in D, but now the exposure of interest is gene A function which is proxied by the variant effect on downstream genes C−F. Horizontal pleiotropy can be avoided in the absence of cell-type specific regulatory effects (panel A) or in the absence of local co-regulation in the *cis* region (panel B). G - genetic instruments; U - unmeasured confounders.

However, inferences from MR studies are only valid if certain assumptions are met (Burgess et al., 2019; Skrivankova et al., 2021). A key assumption of MR is that the genetic variants are associated with the outcome only via the exposure of interest (Reed et al., 2024). This assumption can be violated by *horizontal pleiotropy*, where the causal effect of the genetic variants on the outcome is mediated by another trait not included in the analysis (Sanderson et al., 2024). Importantly, genetic instruments identified for high-throughput protein, transcript or metabolite measurements are often subject to horizontal pleiotropy, leading to incorrect or misleading MR inferences (Karjalainen et al., 2024; Richardson et al., 2022; Smith et al., 2022) (Figure 1). As an example, Karjalainen *et al*. reported that MR between acetone and 233 other metabolites identified 20 significant associations, mostly with lipid traits, but almost all these associations were attenuated when pleiotropic variants at well-known lipid loci were excluded (Karjalainen et al., 2024). Restricting the analysis to four less pleiotropic instruments identified a putative causal association between plasma acetone level and hypertension (Karjalainen et al., 2024). Similarly, both proteomic and transcriptomic studies have identified pleiotropic regulatory variants associated with the abundance of tens to hundreds of genes or proteins (Freimann et al., 2024; Sun et al., 2023; Võsa et al., 2021), reflecting a high degree of co-regulation in *trans*-QTL networks (Figure 1C).

To avoid these pleiotropic effects, many studies focus on *cis*-acting genetic variation to identify the putative causal effect of drug target (typically a gene or protein) perturbation on the outcome of interest (Figure 1A). This approach is referred to as *cis*-MR (Figure 1B). In *cis*-MR, gene expression or protein abundance in an accessible tissue is typically used as an exposure. However, *cis*-MR can still be subject to two types of horizontal pleiotropy. First, if the gene or protein affects the outcome in one cell type or developmental stage but is measured in another one then this can lead to overdispersion heterogeneity or allelic spread that can bias the MR estimates (Patel et al., 2023; Tambets, Kolde, et al., 2024) (Figure 1A). Fortunately, a number of pleiotropy-robust MR methods have been developed to address this (Tambets, Kolde, et al., 2024; van der Graaf et al., 2024; Zhu et al., 2021). Secondly, *cis*-MR can also be subject to co-regulation between neighbouring genes (Figure 1B) (Tambets, Kolde, et al., 2024). Despite these limitations, *cis*-MR has been successfully used to identify known causal relationships in multiple benchmarks (Karim et al., 2023; Porcu et al., 2019; van der Graaf et al., 2024; Zheng et al., 2020). However, current transcriptomic datasets are limited in sample size for most cell types and tissues (Kerimov et al., 2023; Tambets, Kolde, et al., 2024; The GTEx Consortium, 2020) and plasma proteomic studies with large sample sizes only cover a subset of the proteome (e.g. 2,923 proteins in the UK Biobank (Sun et al., 2023)).

Importantly, the variant effect on gene or protein function can also be captured by its effect on proximal downstream phenotypes in metabolic pathways or regulatory networks (Figure 1E). For example, for well-known lipid loci, recent *cis*-MR studies have used variant effect on plasma LDL cholesterol level as a proxy measure for variant effect on protein function (Richardson et al., 2022; Yang et al., 2024). *Cis*-MR where the exposure is a metabolite or another biomarker is sometimes also referred to as drug target MR (Richardson et al., 2022). Similarly, we have used downstream *trans*-eQTL effects to characterise the impact of a lupus-associated *USP18* missense variant on its protein function (Freimann et al., 2024). However, a systematic analysis of when and how these proxy measures for gene or protein function can be used for causal inference is still lacking.

In this study, we expand on the use of high-throughput plasma metabolite measurements as proxy measures for protein function in the *cis*-MR framework. Using genotype and nuclear magnetic resonance (NMR) spectroscopy data from 246,683 UK Biobank participants, we identify 107 confidently fine mapped missense variants for 56 metabolites. In two case studies involving glycolysis and vitamin D synthesis pathways, we demonstrate how the missense variants’ effects on pyruvate and histidine levels can be used as proxy readouts for their effect on *cis* protein function, allowing us to infer causal relationships between disruption of protein function and downstream traits. Finally, we propose a theoretical framework that outlines the key assumptions that need to be satisfied to generalise this approach to other proteins and traits.

## Results

We performed GWAS and fine mapping for 56 metabolites in the UK Biobank using the nuclear magnetic resonance (NMR) platform from Nightingale Health (see Methods). The analysis included 246,683 individuals of European ancestries (see Methods). In total, we identified 107 confidently fine mapped (posterior inclusion probability (PIP) > 0.8) missense variants that were associated with one or more metabolites. All summary statistics and fine mapping results are publicly available (see Data availability). Below, we will present two case studies: one focusing on the effect of glycolysis pathway activity on red blood cell count and another one exploring the role of histidine ammonia lyase (*HAL*) in modulating vitamin D levels.

### Glycolysis pathway, plasma pyruvate level and red blood cell count

Loss-of-function mutations in the pyruvate kinase L/R (*PKLR)* gene are the most common cause of haemolytic anaemia, a disorder in which red blood cells are destroyed faster than they are made (Zanella et al., 2007). In our analysis, we identified eight missense variants (including two variants in the *PKLR* gene) that were robustly associated with plasma pyruvate level (Figure 2A). Reassuringly, the two *PKLR* variants (1_155291845_C_T, rs113403872, and 1_155291918_G_A, rs116100695) were also associated with red blood cell count (RBC), thus confirming the known disease association (Figure 2A) (Zanella et al., 2007). Despite the strong association at the *PKLR* locus, there is no obvious causal mechanism directly linking levels of circulating pyruvate to RBC counts. However, when performing MR between plasma pyruvate level and RBC count using these eight missense variants as instruments, we detected a non-zero “causal” effect (Figure 2B). Notably, there was considerable heterogeneity among the causal effect estimates (Wald ratio) provided by individual genetic instruments, prompting further investigation.

**Figure 2.**
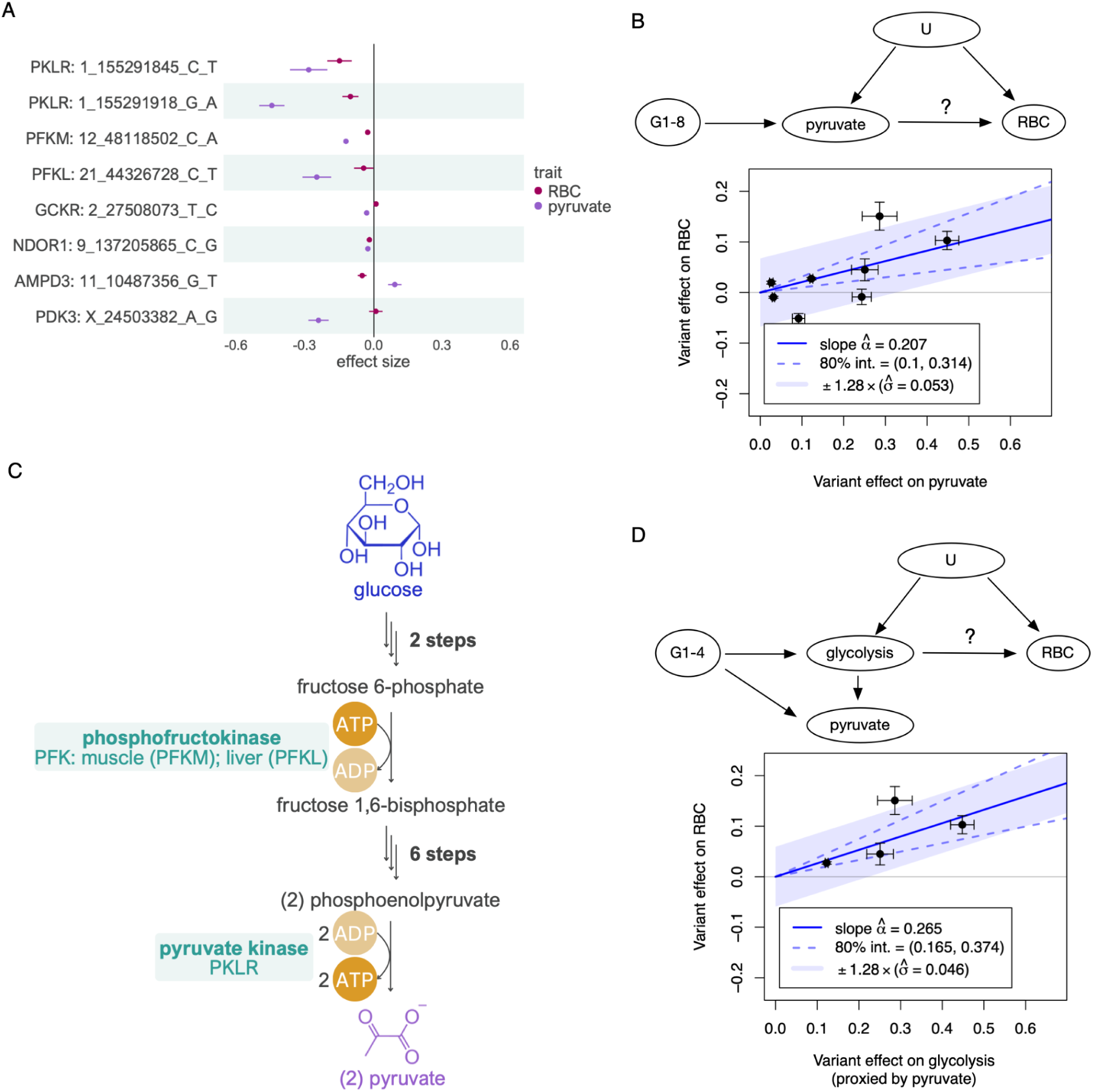
Relationship between plasma pyruvate level and red blood cell count. (**A**) Eight fine mapped missense variants associated with plasma pyruvate level and their effect on red blood cell (RBC) count. (**B**) Mendelian randomisation between plasma pyruvate level (exposure) and RBC count (outcome) using all eight fine mapped missense variants as instruments. (**C**) Role of phosphofructokinase (encoded by *PFKM* and *PFKL* genes) and pyruvate kinase (encoded by *PKLR*) in the glycolysis pathway. Complete pathway is shown in Figure S1. (**D**) Mendelian randomisation between glycolysis pathway activity (exposure) and RBC count (outcome), restricted to missense variants in the three genes (*PKLR*, *PFKM* and *PFKL*) that encode enzymes involved in the glycolysis pathway. The causal diagram illustrates how plasma pyruvate level acts as a proxy for glycolysis pathway activity in red blood cells. G - genetic instruments; U - unmeasured confounders.

We noticed that in addition to the two *PKLR* missense variants, two more missense variants affected another core enzyme of the glycolysis pathway (12_48118502_C_A, rs4760682 in *PFKM* and 21_44326728_C_T, rs118106526 in *PFKL*, both encoding the phosphofructokinase enzyme) (Figure 2C, Figure S1). Given that mature red blood cells lack both nuclei and mitochondria, their energy production, which is essential for their survival, relies entirely on the glycolysis pathway (van Wijk & van Solinge, 2005). As the end product of the glycolysis pathway is pyruvate (Figure 2C), we hypothesised that for these four missense variants, plasma pyruvate level might serve as a proxy readout for the glycolysis pathway activity in RBCs (see causal diagram on Figure 2D).

Indeed, for the four missense variants in the *PFKM*, *PFKL* and *PKLR* genes, we observed directionally concordant effects between reduced plasma pyruvate level and decreased RBC count (Figure 2A). This was further supported by Mendelian randomisation, which now showed considerably better concordance between the causal effect size (Wald ratio) estimates provided by the individual variants (Figure 2D). Importantly, we were now seeking to infer the effect of glycolysis pathway activity on RBC count, rather than the effect of circulating pyruvate levels. Hence, we are using the variants’ effects on plasma pyruvate level only as a proxy to capture their effects on glycolysis pathway activity in RBCs.

As a final validation, we repeated the MR analysis using the four missense variants in genes that do not encode enzymes directly involved in the glycolysis pathway (*GCKR*, *NDOR1*, *AMPD3*, *PDK3*) and detected a null effect (Figure S2), indicating that the initial genome-wide MR estimate (Figure 2B) was primarily driven by the missense variants in genes encoding enzymes of the glycolysis pathway. Notably, the glucokinase regulator (*GCKR*) missense variant (rs1260326, *GCKR*:p.Leu446Pro) is a highly pleiotropic locus associated with 51 (out of 56) selected metabolites in our recent meta-analysis of 599,249 individuals (Tambets, Kronberg, et al., 2024). This example highlights how the levels of plasma metabolites can be regulated through multiple distinct mechanisms. However, even if the metabolite itself (e.g. pyruvate) is unlikely to have a direct causal effect on the outcome of interest (RBC count), it can still act in a locus-specific manner as a proxy measure for other biological traits (e.g. glycolysis) that do have a causal effect.

### Histidine, UV exposure and vitamin D

We recently noticed an interesting common variant (MAF = 42%) GWAS hit near the *HAL* gene (12_95984993_C_T, rs3819817) that was associated both with vitamin D levels (Manousaki et al., 2020) and skin cancer (Seviiri et al., 2022). In the Open Targets Genetics portal (Mountjoy et al., 2021), this variant was identified as an eQTL for the *HAL* gene and was also associated with *trans*-urocanate level in urine (Schlosser et al., 2020) and childhood sunburn occasions (Neale Lab), but was not pleiotropically associated with any other disease (Figure 3A-B). Furthermore, the lead variant had a positive effect on *HAL* expression and *trans*-urocanate level and a negative effect on vitamin D level, sunburn occurrences and skin cancer risk (Figure 3C).

**Figure 3.**
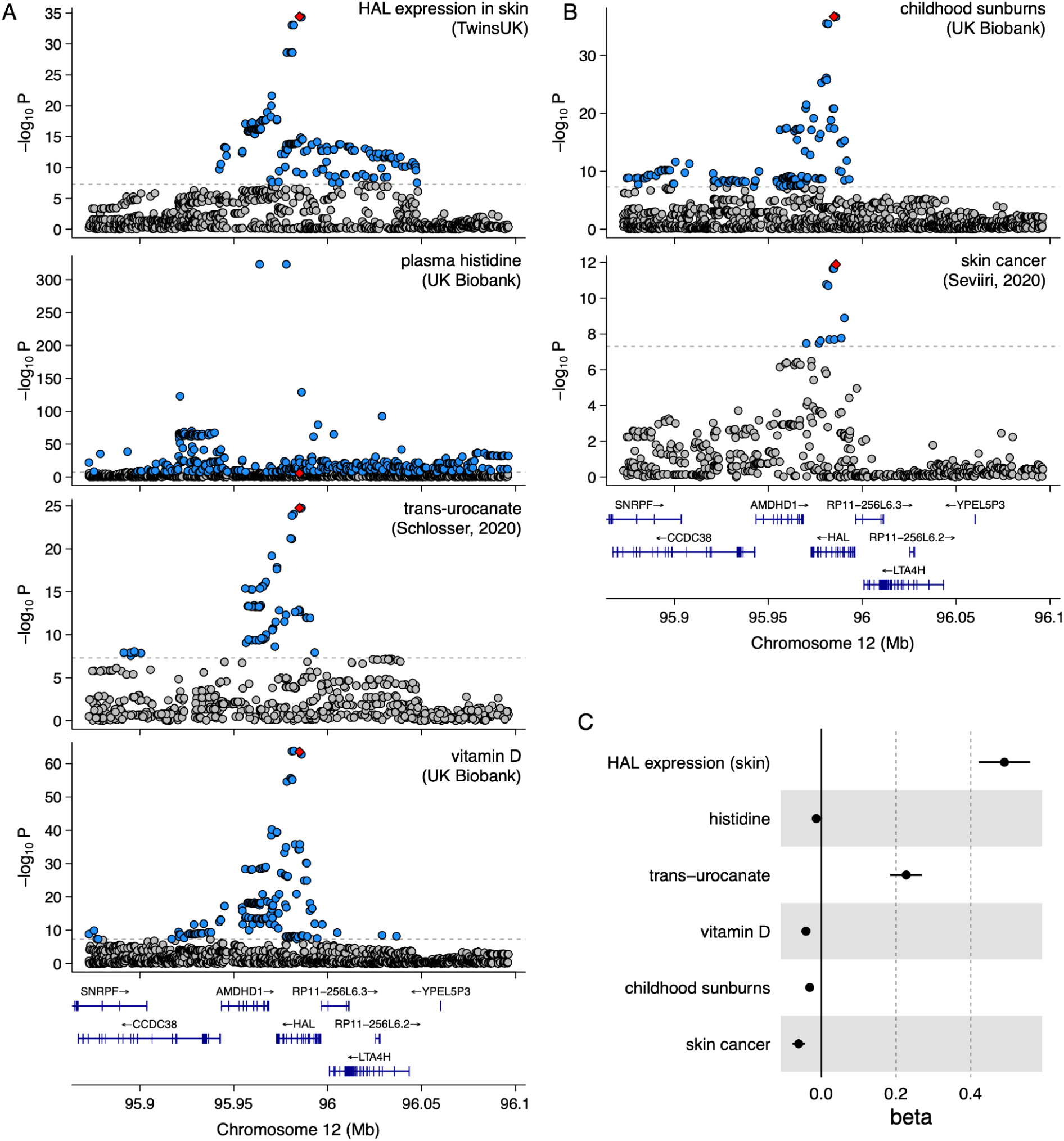
Effect of skin-specific regulatory variation on vitamin D level and other related traits. **(A)** Regional association plots for *HAL* expression in skin, plasma histidine, *trans*-urocanate, and vitamin D levels, illustrating statistically significant associations within the same genomic region. The lead *HAL* eQTL variant (rs3819817) has been highlighted in red. (**B**) Regional association plots for childhood sunburns and skin cancer in the same genomic region. (**C**) Effect size of the rs3819817 *HAL* eQTL lead variant on the six traits.

The biochemical role of histidine ammonia lyase (HAL) in regulating vitamin D levels is well understood (Figure 4). HAL is an enzyme that converts histidine to *trans*-urocanate (Hall, 1952). As a natural sunscreen, *trans*-urocanate absorbs UV light and isomerises to its *cis*-form. This process reduces the effective UV radiation dose in humans, thereby inhibiting vitamin D synthesis. However, the lower dose may also provide protection against sunburn and skin cancer (Barresi et al., 2011). In the liver, *trans*-urocanate is further converted by the urocanate hydratase (encoded by *UROC1*) into 4-imidiazolone-5-propionate (Figure 4) (Kessler et al., 2004). Interestingly, this conversion does not occur in the skin, as *UROC1* is highly expressed in the liver (median TPM = 75.6 in GTEx) but not in the skin (median TPM = 0.01) (The GTEx Consortium, 2020) leading to *trans*-urocanate accumulation in the skin and thus to reduced vitamin D levels but also protection from skin cancer (Figure 3C).

**Figure 4.**
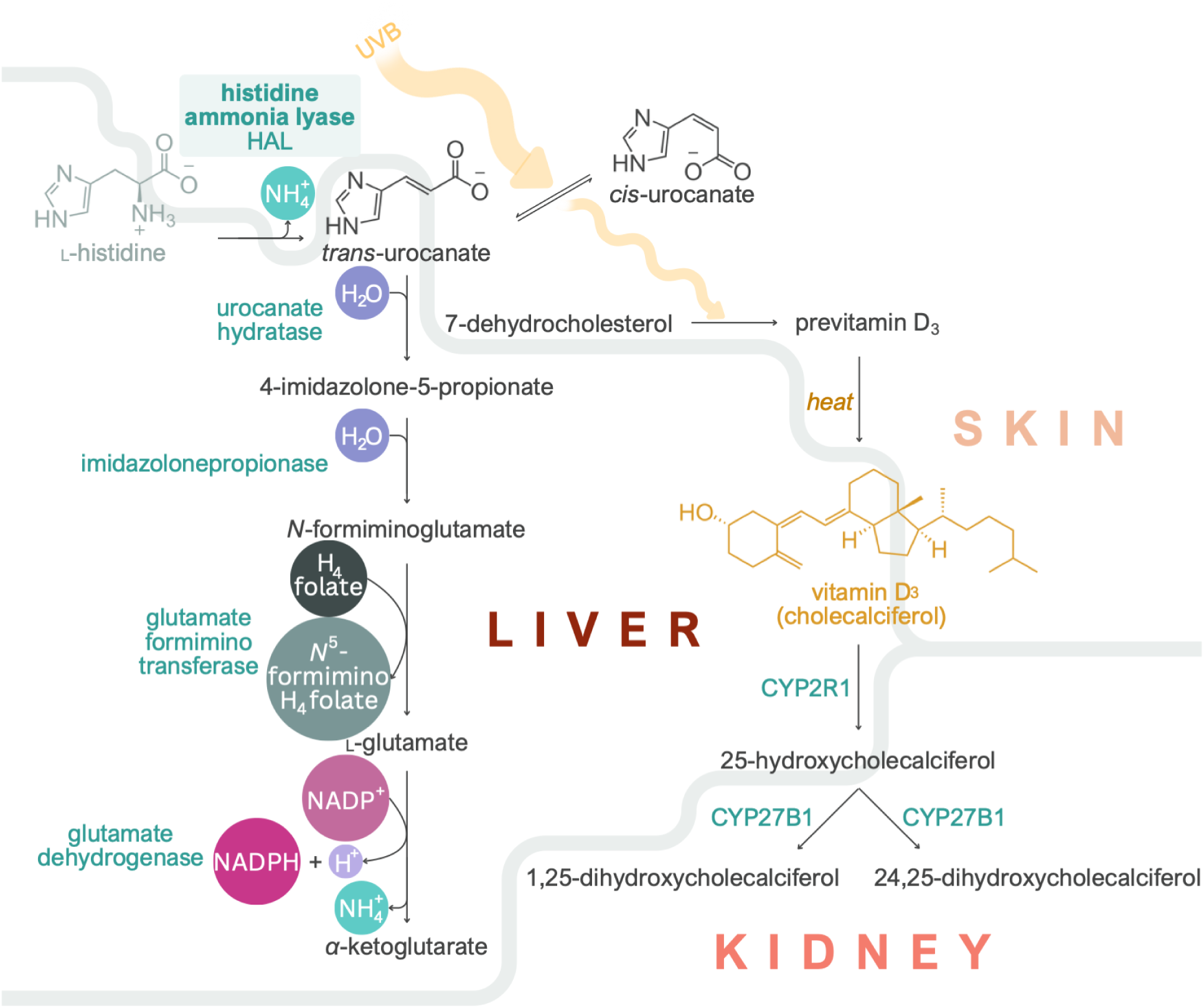
The role of histidine metabolism in regulating vitamin D levels. Role of HAL in regulating vitamin D level in a skin-specific manner.

Unexpectedly, although we anticipated that the effect of the rs3819817 *HAL* eQTL variant on vitamin D level and skin cancer risk would be mediated by the conversion of histidine to *trans*-urocanate (Figure 4), the variant had only a weak association with plasma histidine level (beta = −0.014, *p* = 1.8×10^−6^, Figure 3). To understand this discrepancy, we examined the rs3819817 *HAL* eQTL effect sizes and *p*-values in all 127 datasets in eQTL Catalogue release 6 (Kerimov et al., 2023). The eQTL was highly tissue-specific and detected only in three skin datasets from the TwinsUK (Buil et al., 2015) and GTEx (The GTEx Consortium, 2020) studies (Figure S3). This suggests that the rs3819817 *HAL* eQTL variant primarily affects histidine level in the skin rather than in plasma, via tissue-specific regulation of *HAL* gene expression.

Since histidine was one of the 56 metabolites profiled in our analysis, we next focussed on fine mapped missense variants associated with plasma histidine. Reassuringly, two of the strongest associations corresponded to two low-frequency (MAF < 0.5%) missense variants in the *HAL* gene (12_95977953_C_T, rs61937878, and 12_95986106_C_T, rs117991621), which were also strongly associated with vitamin D levels (Figure 5A) (Kanai et al., 2021). Interestingly, a third missense variant (12_95994812_G_A, rs143854097) in the *HAL* gene was only associated with histidine level and not with vitamin D level, potentially due to its low allele frequency (MAF ∼ 0.1%) and limited statistical power. We also detected missense variants in further seven genes (including the pleiotropic *GCKR*:p.Leu446Pro missense variant also associated with pyruvate) that were robustly associated with histidine but not vitamin D levels, suggesting that plasma histidine is unlikely to have a direct causal effect on vitamin D levels. Unfortunately, we were not able to assess the effects of the three fine mapped *HAL* missense variants on skin cancer, *trans*-urocanate and *HAL* expression due to the low allele frequency of these variants (MAF < 0.5%).

**Figure 5.**
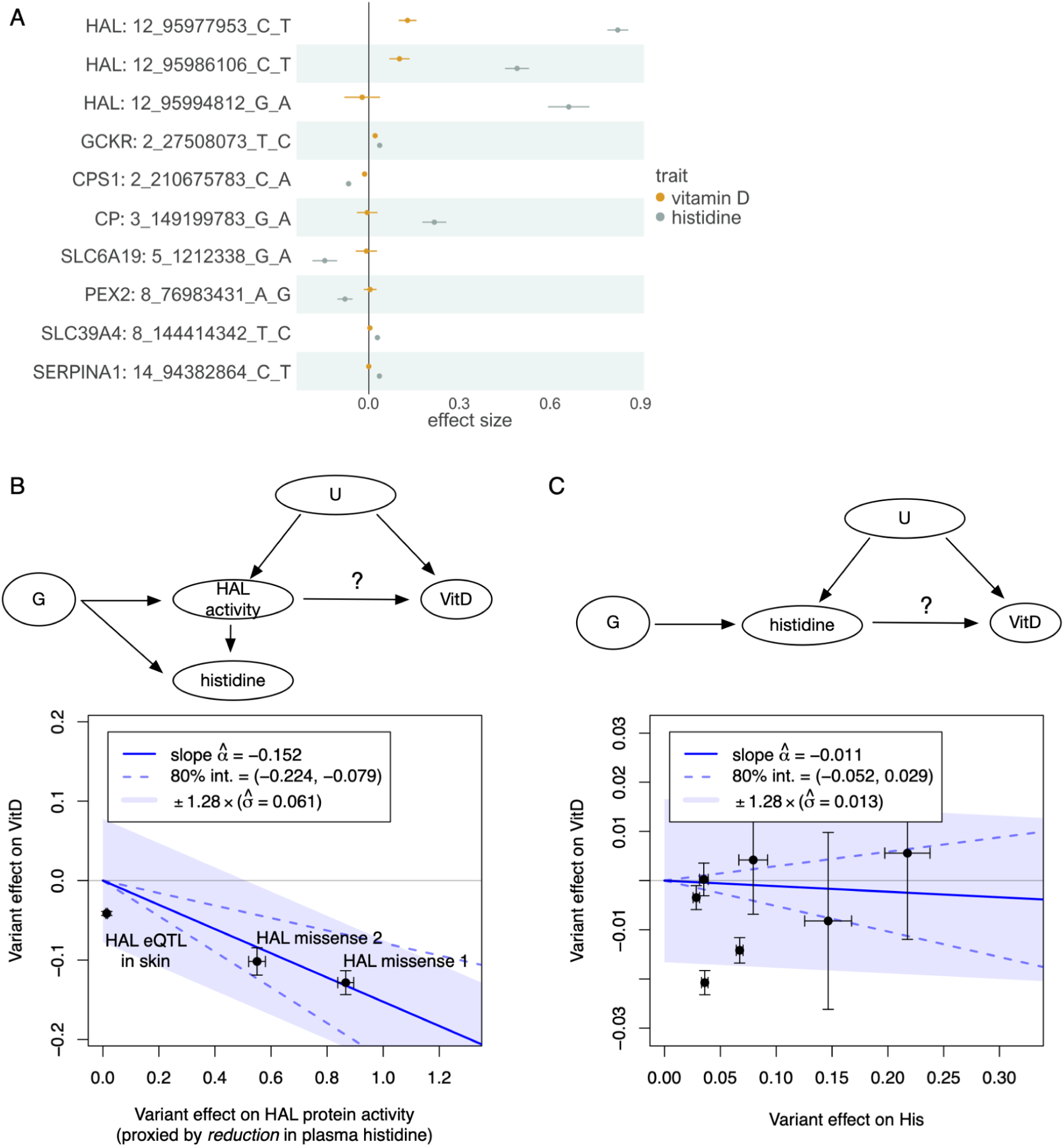
Using plasma histidine level as a proxy for HAL protein activity. (**A**) Fine mapped (PIP > 0.8) missense variants associated with plasma histidine level in the UK Biobank. (**B**) Mendelian randomisation (MR) analysis examining the relationship between proxied HAL protein activity and vitamin D level. The instruments are restricted to the two missense variants in the *HAL* gene and a skin-specific eQTL for *HAL* (Figure 4A). Here, we use the effect of these variants on reducing plasma histidine level as a proxy measure for their effect on HAL function. (**C**) MR between plasma histidine and vitamin D levels using all fine mapped missense variants associated with plasma histidine levels outside the *HAL* region as instruments. G - genetic instruments; U - unmeasured confounders.

Finally, we hypothesised that for variants affecting the *HAL* gene, we could use their effect on reducing plasma histidine level as a proxy measure for their effect on HAL protein function. Using this approach with MR, we detected a significant causal effect of −0.152 between increased HAL protein function (proxied by reduction in plasma histidine) and vitamin D level (Figure 5B). Notably, this estimate was dominated by the two large-effect missense variants in the *HAL* gene. Using the skin-specific eQTL variant (rs3819817) with plasma histidine level as exposure would have yielded a highly misleading estimate of −0.0414/0.014 = −2.96 (Wald ratio) (Figure 5B). This is because this variant likely has a much larger effect on HAL function in the skin, the causal tissue for vitamin D level, than in the tissues that determine histidine level in plasma. Interestingly, using the expression level of *HAL* in the skin as the exposure (instead of plasma histidine level) with the same instrument yielded a causal effect estimate of −0.0414/0.49 = −0.084 (Wald ratio), which aligns more closely with the estimate from the two missense variants (−0.152). The necessity of considering tissue- and cell type-specific effects poses a significant limitation to using variant effects on circulating metabolites (or other molecular traits) as proxy measures for protein function, as we will discuss in detail below.

As a negative control, we performed MR between plasma histidine and vitamin D levels utilising all fine mapped missense variants associated with plasma histidine levels outside of the *HAL* gene as instruments. This allowed us to directly estimate the causal effect of increasing plasma histidine levels on vitamin D levels (Figure 5B). As expected, we observed a null effect, further reinforcing that vitamin D level is primarily influenced by the HAL enzymatic activity in the skin.

### Additional assumptions of MR with proxy exposures

Both the glycolysis and vitamin D examples illustrate how restricting genetic instruments to specific gene regions and using variant effects on plasma metabolites as proxy measures for corresponding gene function can help reduce horizontal pleiotropy and infer plausible causal relationships between perturbed gene function and outcomes of interest. However, generalising this approach to other gene regions and potential proxy exposures requires careful consideration of two key assumptions:

1. **The instruments (genetic variants) must be unambiguously linked to the causal *cis*-gene**. The majority of trait-associated genetic variation is non-coding, likely modulating the expression or splicing of nearby *cis* genes. We and others have shown that expression-altering variants often regulate the expression of multiple neighbouring genes (Tambets, Kolde, et al., 2024) (Figure 1B). Although splicing QTLs tend to have more specific effects on a single target gene, distinguishing them from expression QTLs can be challenging in practice (Kerimov et al., 2023). This is the main reason why we focused on fine mapped missense variants in this study, as they can be linked to the causal gene with high confidence. However, missense variants are rare and may not be available for most traits and exposures. Thus potential violation of this assumption should be explicitly considered when performing analyses such as drug target MR that include all genetic variants from a specific gene region as instruments (Gill et al., 2024; Richardson et al., 2022; Yang et al., 2024).
2. **For accurate inference, the proxy metabolite, transcript, or protein being measured should be *downstream* and *proximal* to the *cis*-gene or protein of interest whose function we are aiming to approximate.** In our case study, for variants affecting the *HAL* gene, it is preferable to use histidine or *trans*-urocanate concentrations rather than metabolites further downstream in the pathway (Figure 3B). In practice, however, the exact mechanisms by which the *cis* gene affects the measured traits are often unclear, which could inadvertently result in capturing traits that are downstream of the outcome of interest, potentially leading to reverse causation. As GWAS sample sizes increase, the proportion of discoveries that correspond to these indirect effects is also likely to increase. For example, in a very large meta-analysis of NMR metabolites (n = 599,249), the *HAL* missense variant rs61937878 was also weakly associated the plasma glycine levels (beta = 0.071; p = 2.5x10^-10^), likely reflecting an indirect pleiotropic effect (Figure S4).

In addition to these two assumptions specific to proxy exposures, we also need to consider the factors that can invalidate any *cis*-MR analysis with molecular traits as exposures. First, molecular traits such as gene expression, protein abundance or metabolite concentrations can often be measured in many different cell types, tissues or developmental stages (contexts for short). In an ideal scenario, the context in which the genetic variant’s effect on the exposure has a causal effect on the outcome (‘causal context’) is the same where the exposure is measured (‘proxy context’), but this is often not the case. In the *HAL* example, the likely causal context where *HAL* influences vitamin D levels is skin tissue, but the proxy context in which histidine was measured is plasma. If a genetic variant has the same effect on the exposure in the proxy context as it would in the causal context (e.g. it is a missense variant), then context misspecification is less important. However, non-coding regulatory variants can often have context-specific effects, and this can significantly bias MR estimates. For example, the skin-specific eQTL for *HAL* had almost no effect on plasma histidine level (Figure 3A). Secondly, even if the included instruments themselves do not have context-specific effects, they might still be in LD with other context-specific genetic variants that do. This can bias the marginal effect sizes of the instruments on the exposure, thus also biasing the MR estimates when using an exposure measured in the proxy context. A promising approach to account for these biases are methods such as MR-link-2 that explicitly model the LD between instruments and their potentially pleiotropic effects (van der Graaf et al., 2024).

## Discussion

Using examples from the glycolysis and vitamin D synthesis pathways, we have constructed two case studies to demonstrate how horizontal pleiotropy can mislead MR to infer implausible causal relationships between an exposure and an outcome. Our case studies complement previous reports highlighting widespread horizontal pleiotropy affecting plasma metabolite levels and other high-throughput molecular measurements (Freimann et al., 2024; Karjalainen et al., 2024; Richardson et al., 2022; Smith et al., 2022; Yang et al., 2024). We illustrate how MR analysis can be reformulated by focusing on genetic variation located in *cis* of specific target genes and using the high-throughput molecular measurements as proxy readouts of protein function (Figure 1E). The key contribution of our work is to explicitly outline the additional assumptions required for this approach to produce valid inferences. We expand on previous work focused on well-known lipid loci (Richardson et al., 2022; Yang et al., 2024) by providing a general framework for conducting MR analysis using arbitrary proxy measures of protein function.

A related ‘*trans*-weighted *cis*-MR’ idea was presented in the MR-Fish study (Warwick et al., 2024). However, a key difference between our analysis and theirs is that they did not explicitly consider the assumptions that the instruments and proxy exposures should satisfy to produce reliable inferences. For instance, by using variants in the *FTO* locus as instruments and plasma CRP level as the proxy exposure, the authors inferred a putative causal link between altered *FTO* function (proxied by variant effect on CRP) and type 2 diabetes risk. However, they overlooked that the lead non-coding variant at the *FTO* locus regulates the expression of *IRX3* and *IRX5* transcription factors instead of the *FTO* gene itself (Claussnitzer et al., 2015), thereby violating our first assumption. Thus, it is unclear what is the added value of the MR Fish approach beyond simply reporting the closest genes at the outcome-associated locus, because there is no guarantee that the included instruments have any effect on the claimed *cis* gene. They also did not consider our “proximal effect” assumption (assumption 2), which could easily lead to cases of reverse causation, where the variant effect on the exposure is mediated via the outcome.

Most *cis*-MR analyses use gene expression levels or protein abundances as exposures (Karim et al., 2023; Porcu et al., 2019; Tambets, Kolde, et al., 2024; van der Graaf et al., 2020; Zheng et al., 2020). However, when the aim is to infer the causal relationship between altered protein function and an outcome of interest, gene expression or protein abundance are themselves imperfect proxies of protein function. This could be particularly problematic for missense and splice regulatory variants, as their effect on gene expression and protein abundance might be poorly correlated with protein function due to the existence of distinct functional isoforms (Gotthardt et al., 2023; Park et al., 2018; Wright et al., 2022) or because of assay-specific quantification artefacts (Eldjarn et al., 2023; Pietzner et al., 2021). Using downstream regulatory or metabolic effects as proxy measures for protein function mitigates these limitations. For example, using *HERC5* gene expression in lymphoblastoid cell lines as a readout of the missense variant effect on USP18 protein function allowed us to establish a potentially causal link between reduced USP18 function and increased lupus risk (Freimann et al., 2024). This would not have been feasible using the standard *cis*-MR approach, as the missense variant had no effect on *USP18* gene expression (there was no *cis-*eQTL), and USP18 protein abundance has not been measured in the disease-relevant context.

Using proxy exposures in *cis*-MR also has several limitations, as outlined by the assumptions above. In particular, the variant mechanisms of action for most detected genetic signals are often unknown, making it challenging to unambiguously link the variants to the causal genes. Furthermore, for most metabolite GWAS signals and *trans*-QTL loci, we lack sufficient mechanistic understanding to determine whether the detected effect is proximal or indirectly mediated by other factors. In fact, one of the main reasons we knew to focus on pyruvate and histidine in our two case studies were the names of the two enzymes prompting the analysis: *pyruvate* kinase and *histidine* ammonia lyase. These limitations can restrict the practical utility of using proxy exposures in MR, and we caution against performing automated all-against-all *cis*-MR analyses with proxy exposures without careful consideration of the underlying assumptions.

## Methods

### Study cohort

The UK Biobank is a longitudinal biomedical study of approximately half a million participants between 38-71 years old from the United Kingdom (Bycroft et al., 2018). Participant recruitment was conducted on a volunteer basis and took place between 2006 and 2010. Initial data were collected in 22 different assessment centers throughout Scotland, England, and Wales. Data collection includes elaborate genotype, environmental and lifestyle data. Blood samples were drawn at baseline for all participants, with an average of four hours since the last meal, i.e. generally non-fasting. NMR metabolomic biomarkers (Nightingale Health, quantification library 2020) were measured from EDTA plasma samples (aliquot 3) during 2019–2024 from the entire cohort. Details on the NMR metabolomic measurements in UK Biobank have been described previously for the first tranche of ∼120,000 samples (Julkunen et al., 2023). The UK Biobank study was approved by the North West Multi-Centre Research Ethics Committee. This research was conducted using the UK Biobank Resource under application numbers 91233 and 30418.

### Metabolite measurements

This dataset encompassed both the tranche one dataset, comprising approximately 130,000 samples, and the tranche two dataset, which augmented the resources with an additional 170,000 samples. Details of Nightingale’s NMR metabolomics platform and the biomarker measures have been provided for UK Biobank’s metabolomics Supplier Criteria Tables in July 2016 (project reference 15004). For the current research, 56 biomarkers from the available panel were selected for GWAS analysis and fine mapping (Table S1). We excluded individuals with more than 5 missing metabolite measurements from the cohort and applied a metabolite-wise inverse normal transformation to obtain the final dataset.

### PCA-based genetic ancestry assignment

We performed principal component analysis (PCA) of the genotype data using FlashPCA2 (Abraham et al., 2017). Subsequently, all individuals within the UK Biobank dataset who also had NMR data available were clustered into genetic ancestry groups based on their first three principal components using GaussianMixture() function from the scikit-learn Python module. The number of mixture components was set to four based on empirical analysis. The final dataset, representing the largest PCA cluster corresponding to predominantly European genetic ancestry individuals, comprised 246,683 individuals.

### Association testing and fine mapping

The association testing between genetic variants and 56 metabolites was conducted using the regenie software (Mbatchou et al., 2021). During the analysis, sex and the ten top genotype PCs calculated with FlashPCA2 were utilised as study-specific covariates. In regenie step 1, the linkage disequilibrium (LD) pruned variants were used as an input. LD pruning was performed with PLINK2 with the following parameters: MAF > 0.001, window size = 50000 variants, window shift at the end of each step = 200 variants and pairwise r2 threshold = 0.05. In regenie step 2, the minimum imputation info score was set to 0.6, and the minimum minor allele count was calculated based on the number of samples so that MAF would be equal to 0.001.

After association testing, the statistical fine mapping on the summary statistics obtained from regenie and in-sample LD matrix was conducted using the Sum of Single Effects Model (SuSiE) (Wang et al., 2020). LD matrices were calculated with LDstore2 (Benner et al., 2017) software for each fine mapped region. Fine mapped regions were defined for each genome-wide significant locus (p < 5x10^-8^) by considering a 3 Mb wide window centred around the lead variant. In cases where these regions overlapped but did not exceed a total span of 6 Mb, they were merged into a single region. If the resulting region exceeded this 6 Mb limit, the originally defined regions were recursively reduced until all regions adhered to this size constraint. (If LDstore2 encountered a segmentation fault in the following step, alternative maximum region limits of 4.5 Mb or 3 Mb were employed instead.) Regions containing fewer than 50 variants were omitted from the analysis. Additionally, due to the extensive LD structure in the region, the major histocompatibility complex (MHC) region (chr6:28,477,797-33,448,354) was excluded from fine mapping. In the SuSiE method (Wang et al., 2020), the maximum number of causal variants within a locus was set to 10. Consequently, up to 10 independent 95% credible sets (CS) and posterior inclusion probabilities (PIP) for each variant were computed, utilising the default uniform prior probability of causality.

Association testing and fine mapping was performed on human genome assembly GRCh37. Subsequently, the coordinates of the imputed variants within the fine mapping results were lifted to the GrCh38 build. This transition was accomplished using the ‘liftover()’ function available in the R package MungeSumstats (Murphy et al., 2021). The Nextflow workflow for GWAS analysis and fine mapping is available from GitHub (https://github.com/AlasooLab/reGSusie).

### External summary statistics

The UK Biobank summary statistics and fine mapping results for red blood cell count and vitamin D were downloaded from Google Cloud (<u>link</u>) (Kanai et al., 2021). The GWAS summary statistics for skin cancer from (Seviiri et al., 2022) study were downloaded from the GWAS Catalog (accession GCST90137411). The GWAS summary statistics for “childhood sunburn occasions” (UK Biobank data field 1737) were downloaded from the Neale lab website (*UK_Biobank_GWAS: Overview of the Data QC, Code, and GWAS Summary Output from the 2017 UK Biobank Data Release*, n.d.).

### Software used

Mendelian randomisation was performed with the fitSlope() function from the MRLocus R package version 0.0.26 (Zhu et al., 2021). All forest plots were made with the ggforestplot R package (https://github.com/NightingaleHealth/ggforestplot).

## Data and code availability

The GWAS summary statistics for the 56 metabolites are available from Zenodo (https://doi.org/10.5281/zenodo.13821209). The fine mapped credible sets and log Bayes factors from SuSiE are available from Zenodo (https://doi.org/10.5281/zenodo.13821038). The GWAS and fine mapping Nextflow workflow is available from GitHub (https://github.com/AlasooLab/reGSusie).

## Data Availability

The GWAS summary statistics for the 56 metabolites are available from Zenodo (https://doi.org/10.5281/zenodo.13821209). The fine-mapped credible sets and log Bayes factors from SuSiE are available from Zenodo (https://doi.org/10.5281/zenodo.13821038). The GWAS and fine mapping Nextflow workflow is available from GitHub (https://github.com/AlasooLab/reGSusie).

https://doi.org/10.5281/zenodo.13821038

## Acknowledgements

I.R., K.A, and R.T. were supported by a grant from the Estonian Research Council (grant no PSG415). This research has been conducted using the UK Biobank Resource under application numbers 91233 and 30418. Nightingale Health Plc is acknowledged for early access to the UK Biobank NMR metabolite data. We thank Adriaan van der Graaf and Zoltan Kutalik for helpful comments on the manuscript.

## Author contributions

I.R. performed genome-wide association testing and fine mapping on the UK Biobank data. R.T. perform colocalisation on the summary statistics from the *HAL* locus. E.B.F. initially identified the association at the *HAL* locus and provided biological interpretation. K.A. conceived the study and performed the MR analyses. I.R and K.A. wrote the manuscript with contributions from all authors.

## Supplementary figures

**Figure S1.**
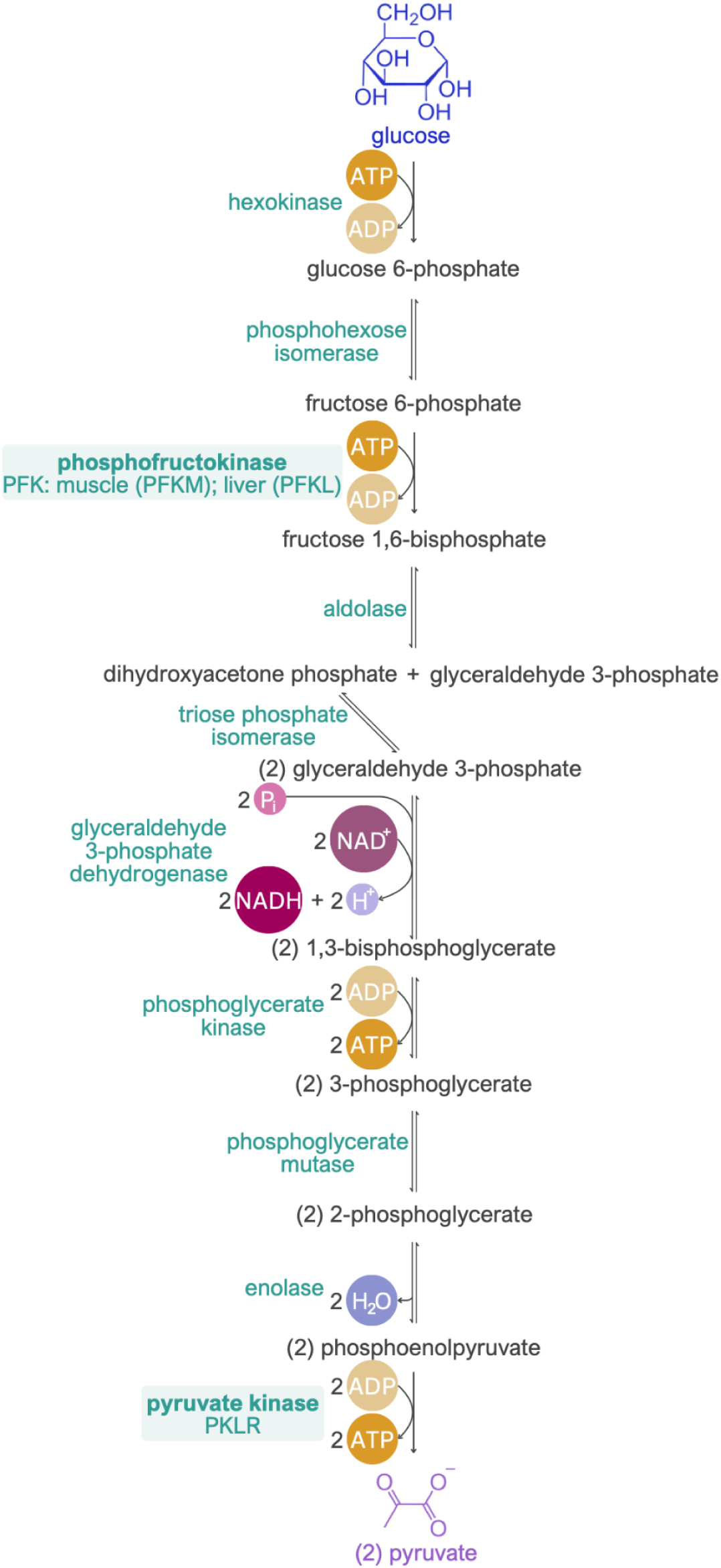
Diagram of the glycolysis pathway.

**Figure S2.**
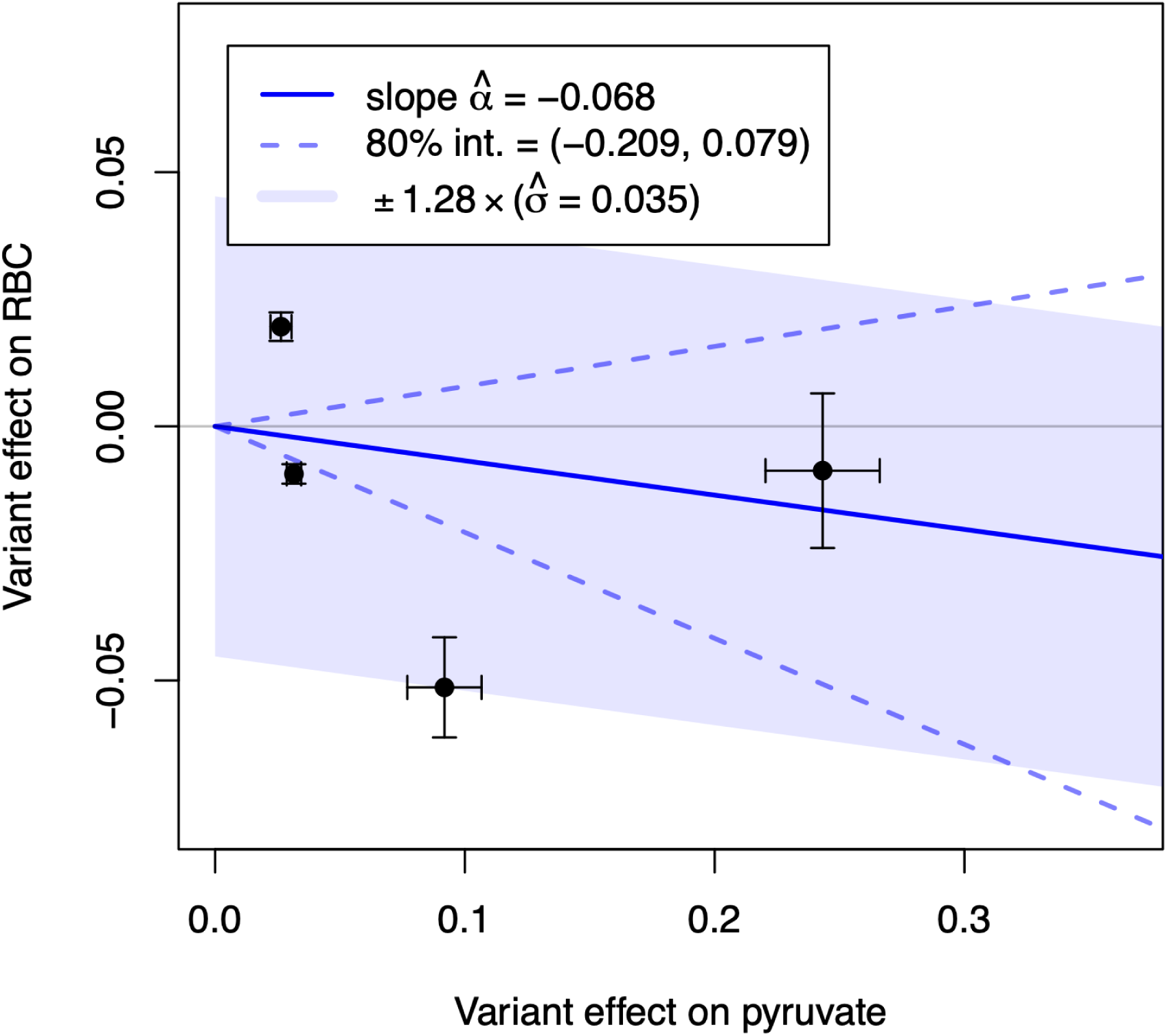
Mendelian randomisation between plasma pyruvate (exposure) and red blood cell count (RBC) (outcome) using missense variants outside of the glycolysis pathway (*GCKR*, *NDOR1*, *AMPD3*, *PDK3*) as instruments.

**Figure S3.**
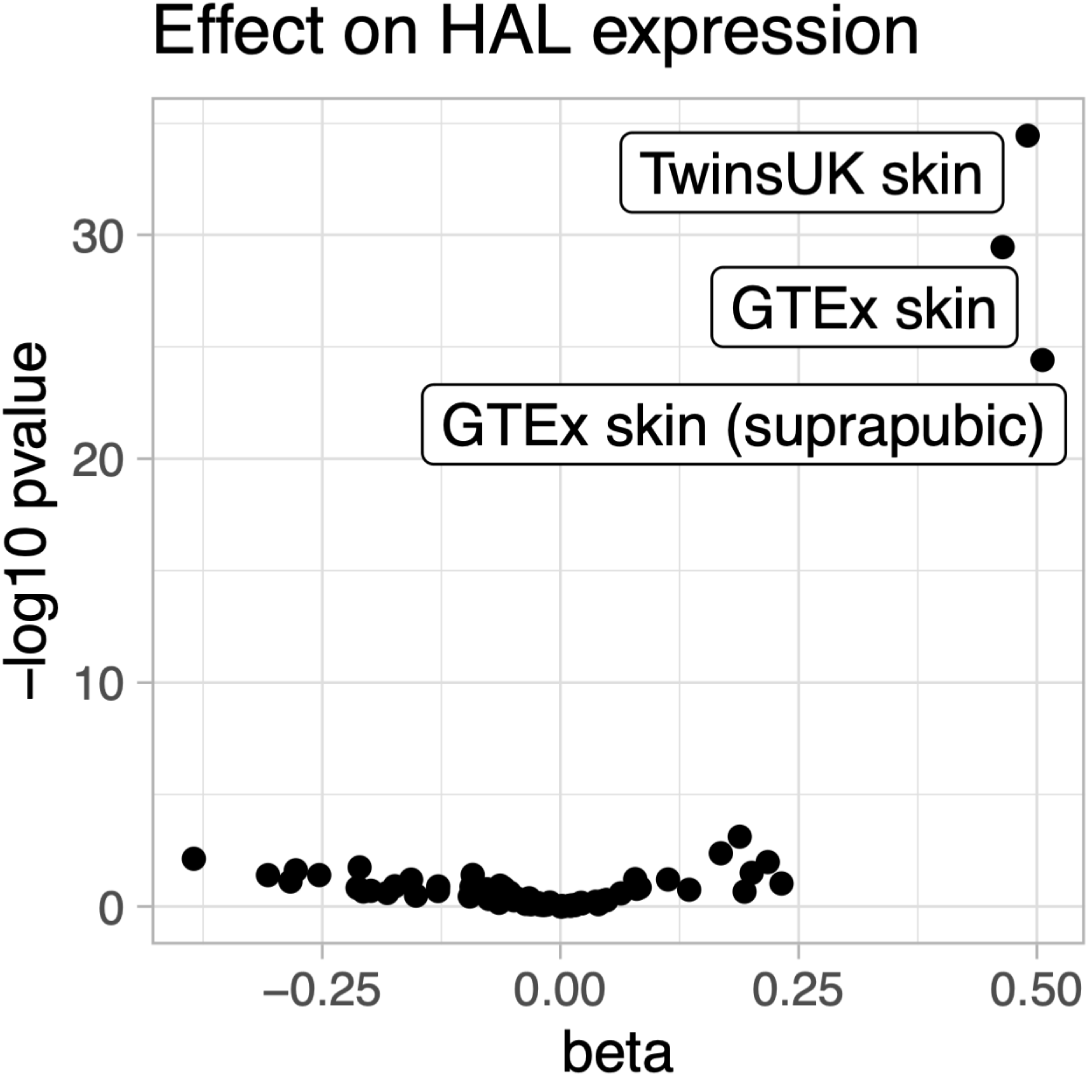
Volcano plot of the vitamin D lead variant effect on *HAL* expression across 127 eQTL Catalogue release 6 datasets.

**Figure S4.**
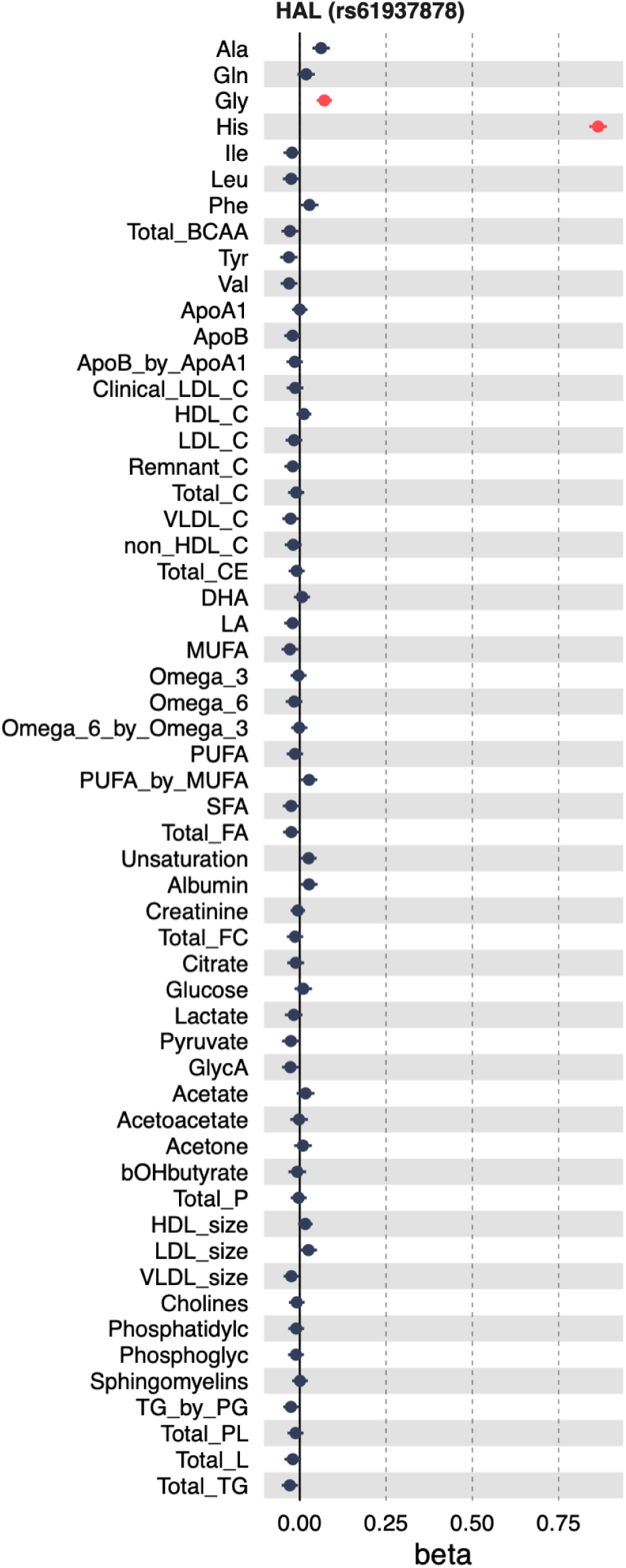
Pleiotropic association between HAL missense variant rs61937878 was plasma glycine levels. The absolute variant effect on glycine (beta = 0.071; p = 2.5x10^-10^) is even smaller than the variant effect on vitamin D (beta = -0.12841), likely reflecting an indirect pleiotropic effect.

